# Detecting and Monitoring Brain Disorders Using Smartphones and Machine Learning

**DOI:** 10.1101/2020.10.03.20206235

**Authors:** Rich Colbaugh, Kristin Glass, Volv Global

## Abstract

The ubiquity of smartphones in modern life suggests the possibility to use them to continuously monitor patients, for instance to detect undiagnosed diseases or track treatment progress. Such data collection and analysis may be especially beneficial to patients with i.) *mental disorders*, as these individuals can experience intermittent symptoms and impaired decision-making, which may impede diagnosis and care-seeking, and ii.) progressive *neurological diseases*, as real-time monitoring could facilitate earlier diagnosis and more effective treatment. This paper presents a new method of leveraging passively-collected smartphone data and machine learning to detect and monitor brain disorders such as depression and Parkinson’s disease. Crucially, the algorithm is able learn accurate, interpretable models from small numbers of labeled examples (i.e., smartphone users for whom sensor data has been gathered *and* disease status has been determined). Predictive modeling is achieved by learning from both real patient data and ‘synthetic’ patients constructed via adversarial learning. The proposed approach is shown to outperform state-of-the-art techniques in experiments involving disparate brain disorders and multiple patient datasets.

## 1. Introduction

The ubiquity of smartphones permits patients to be continuously monitored [1-3], suggesting the possibility to track their health status, detect undiagnosed diseases, evaluate treatment progress, and predict the emergence of new conditions. Such data collection and analysis may be particularly beneficial to individuals suffering from i.) *mental disorders*, as these patients can experience intermittent symptoms, social stigma, and impaired decision-making, which can in turn impede diagnosis and inhibit care-seeking [1,2], and ii.) progressive *neurological diseases*, as real-time monitoring could facilitate earlier diagnosis and more effective and informed treatment design [2,3]. This paper examines the feasibility of using passively-collected smartphone data and machine learning to detect and monitor brain disorders in individuals. To increase generality, it is assumed patients’ smartphones are equipped with only standard sensors: GPS (latitude-longitude coordinates), accelerometers, microphone, ambient light sensor, and screen usage detection.

We focus on depression and Parkinson’s disease as important and illustrative example disorders [e.g. 1-15], but emphasize that the proposed analytic methodology is expected to have broader utility. Depression is a leading cause of disability worldwide, estimated to affect more than 350M people [16]. The DSM [17] lists nine symptoms commonly associated with major depressive disorder: depressed mood, diminished interest and pleasure in activities, fatigue, restlessness, sleep change, weight change, diminished ability to concentrate, feelings of worthlessness, and thoughts of death and suicide. Several of these symptoms appear to have proxies which can be continuously and conveniently inferred from the outputs of sensors available in (standard) smartphones. This observation indicates it may be possible to recognize depression through computational analysis of passively-gathered smartphone data, and indeed this possibility has attracted the attention of clinicians and researchers (see, for example, [1,2,4-9] and references therein).

Parkinson’s disease (PD) is the second most prevalent neurodegenerative disorder worldwide, and incidence is rising [18]. While current treatment strategies aim to improve symptoms, increasing effort is being devoted to developing therapies which can slow or prevent PD progression. Such proactive treatment methods are likely to be most effective when used early in the disease process, before substantial neuronal loss has occurred [19]. However, existing methods for detecting the onset of PD are invasive and expensive, impeding early diagnosis [e.g. 13,18,19]. Symptoms of PD such as motor disfunction and speech impairment may have proxies which can be conveniently and inexpensively measured with smartphone sensors, suggesting the potential to efficiently recognize PD through analysis of smartphone data [10-15].

While promising, proposed smartphone-based methods for detecting and monitoring brain disorders possess several limitations [1-15]. Perhaps most importantly, they suppose requisite models can be induced via conventional statistical or machine learning algorithms [e.g. 20] and thus experience difficulties in the usual situation where only a small amount of labeled training data is obtainable (i.e., there are few individuals for whom target disease status has been determined *and* smartphone data has been collected). Additionally, existing detection/monitoring systems: i.) ordinarily do not base their predictions on clinically-grounded symptoms, which reduces model interpretability, treatment compatibility, and overall clinical relevance, ii.) fail to exploit prior domain knowledge concerning expected relationships between certain behaviors and target disorders (e.g. depression may decrease one’s ability to concentrate), iii.) typically have evaluated model performance with just a single cohort and dataset, making it hard to assess a model’s generalizability and robustness, iv.) often require smartphone data to be augmented with other information (e.g. responses to questionnaires or data from special-purpose sensors).

This paper develops algorithms for learning smartphone-based brain-disorder detection models which overcome the limitations of previous schemes and shows that the learned models outperform state-of-the-art tools. More specifically, we

- propose a framework within which to map smartphone sensor data to clinically-meaningful predictor variables (features), thereby facilitating induction of medically-grounded models;
- introduce a new methodology for learning accurate, scientifically-sound prediction models from small numbers of training examples by:
  - taking supervision from both *instance labels* [20] – individuals labeled as to their disease status – and *feature labels* [21] – tags indicating whether the presence of certain features is expected, according to domain knowledge, to be associated with the target disease;
  - augmenting available (real) patient data with ‘synthetic’ training examples derived via adversarial learning [22] and then using both real and synthetic patients for model training [23];
- demonstrate our learned models outperform both state-of-the-art computational techniques and clinical screening tools for two disparate diseases and across multiple patient datasets.

## 2 Predictive Modeling

### A. Problem Formulation

Suppose we are given access to passively-collected smartphone data for a patient and wish to predict whether the individual has a brain disorder of interest. A supervised learning approach to this task entails: i.) acquiring smartphone sensor traces for a cohort of patients who have been labeled according to whether they have the target disorder (TD), preferably by clinical experts, and ii.) using these examples to learn a model which recognizes the TD-status of new (unseen) persons [20]. Unfortunately, in medical passive-sensing applications there are rarely enough labeled examples to support induction of good, generalizable models.

We propose to construct accurate, interpretable models for detecting target brain disorders, even in settings with few labeled training examples, by using machine learning guided by domain knowledge. The predictive modeling process integrates three steps:

1. *condition-relevant feature mapping* – smartphone sensor readings are mapped to a set of proxies for symptoms and behaviors understood to be related to the target disorder (e.g. depression [17]);
2. *instance + feature label learning* (I+FLL), where instance-label supervision (based on patients with known disease status) is employed to induce a preliminary model, which is then refined through feature label supervision that encodes relevant medical knowledge;
3. *learning from synthetic patients* (LSP), in which available training data is augmented with ‘synthetic’ examples generated via adversarial learning [22], and the resulting dataset is used to train enhanced I+FLL models [23].

The combination of condition-relevant feature mapping and I+FLL forms the ‘baseline’ predictive modeling process (called I+FLL for convenience). LSP is added to this procedure when necessary to overcome the challenges with extremely limited or class-imbalanced training data. The general strategy is depicted in Figure 1, and its instantiation for two important and illustrative TDs, depression and PD, are sketched in Figures 2 and 3.

**Figure 1.**
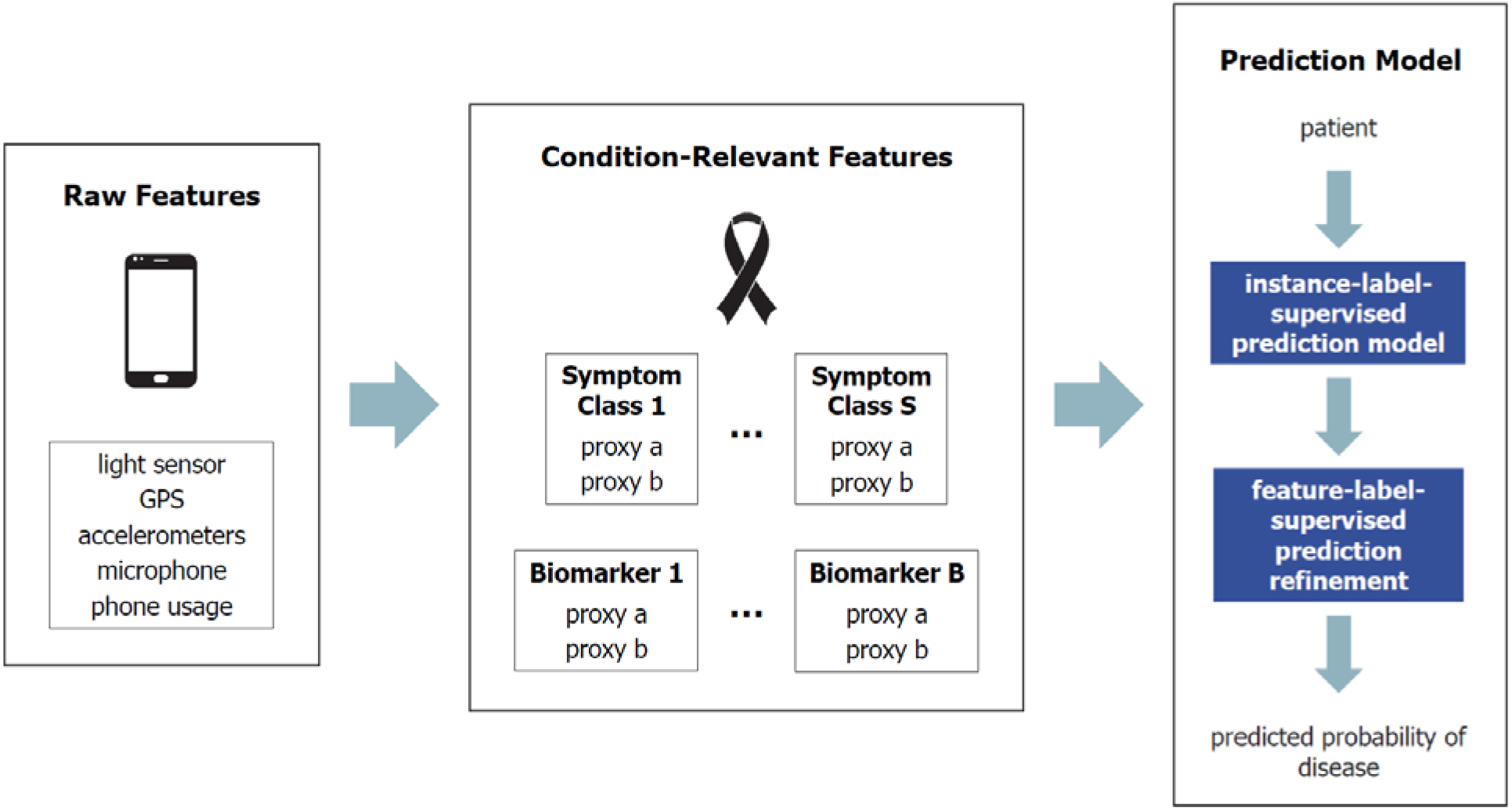
Schematic of general I+FLL process.

**Figure 2.**
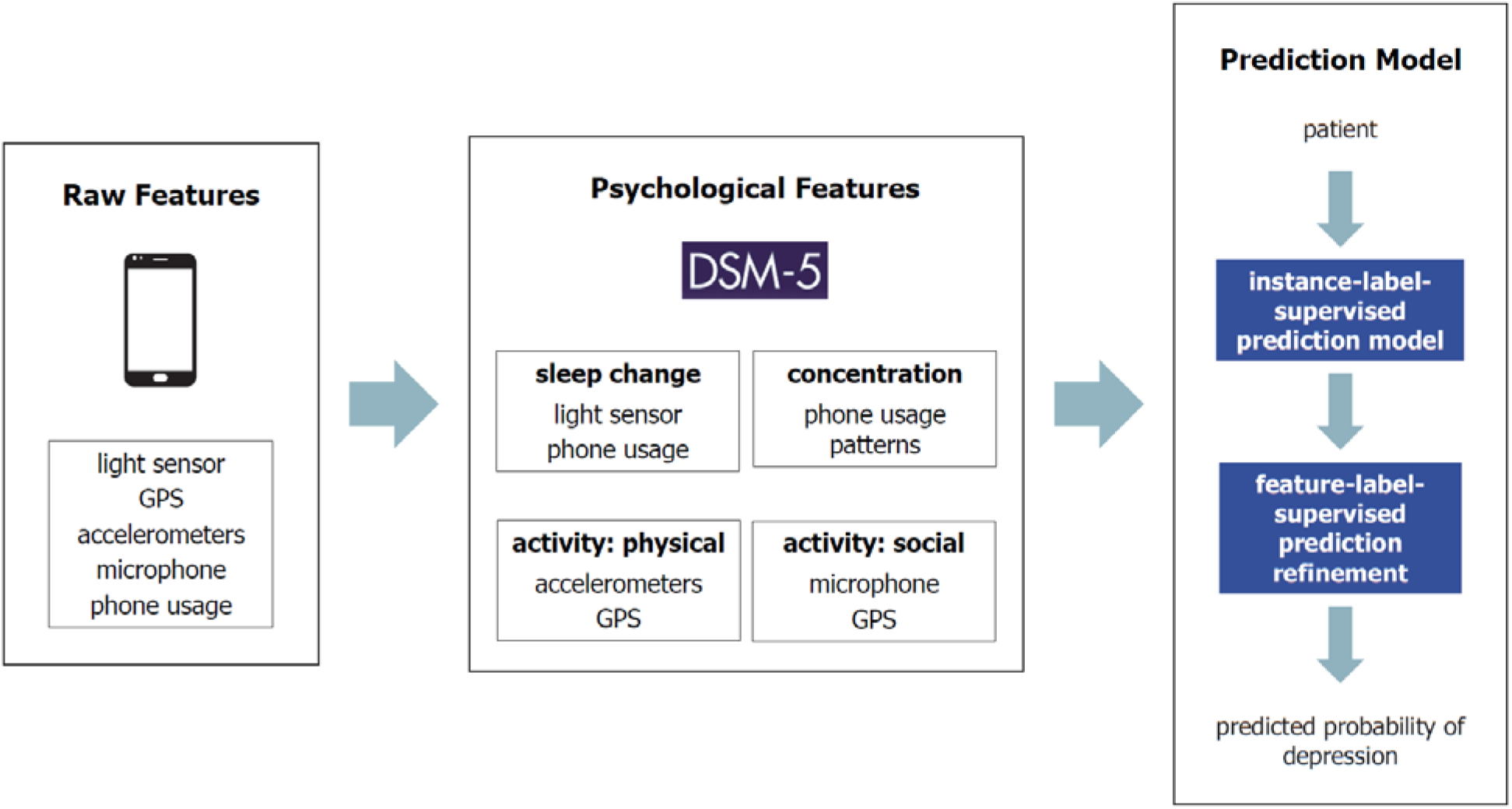
Schematic of I+FLL process for depression.

**Figure 3.**
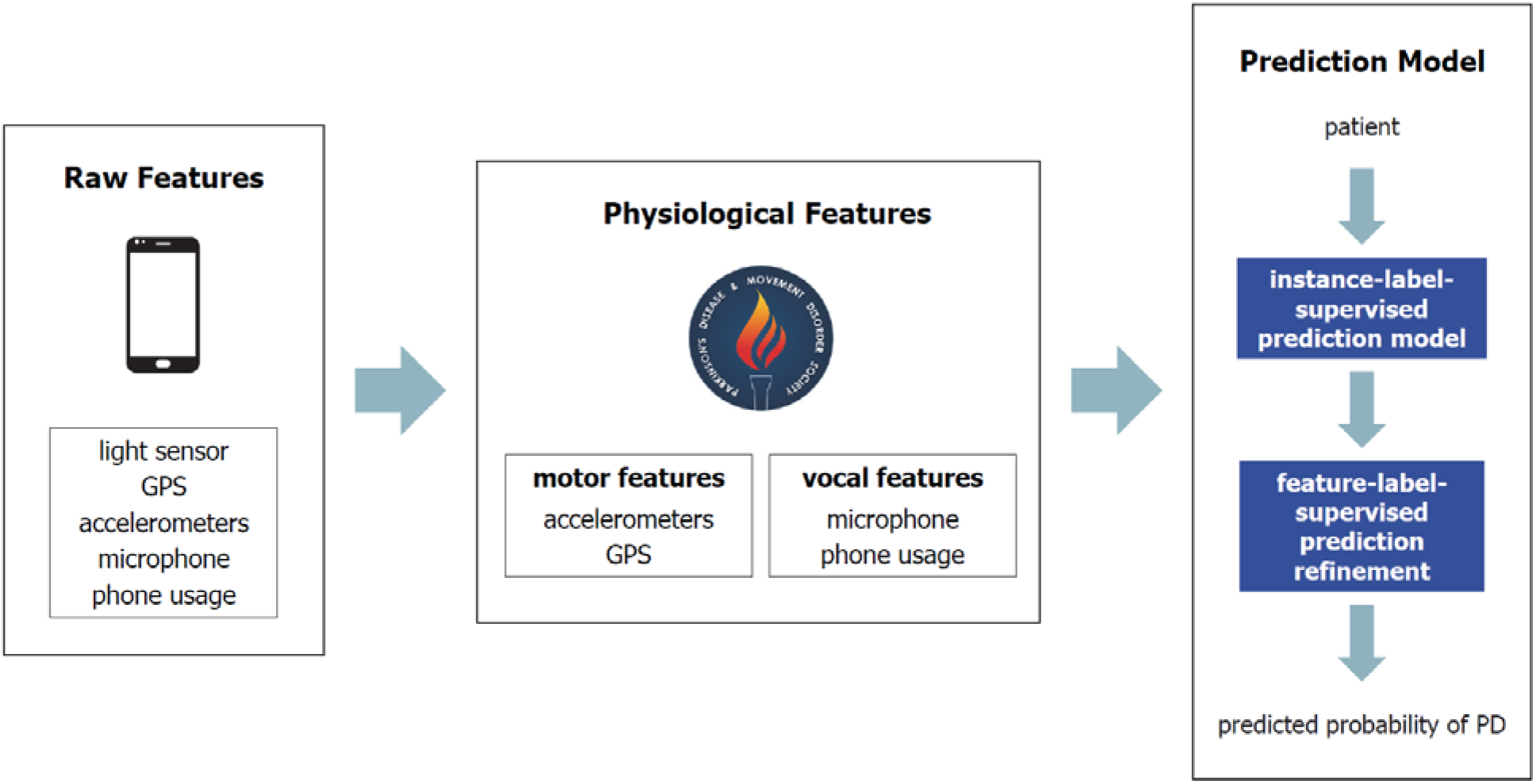
Schematic of I+FLL process for Parkinson’s disease.

At a conceptual level (see Figure 1), predictive modeling begins by mapping raw smartphone sensor outputs to proxies for symptoms and biomarkers associated with the TD. To increase generality, it is assumed that only data measured by standard smartphone sensors (with time-stamps) is available for analysis. These sensors include accelerometers, GPS (latitude-longitude coordinates), microphone, ambient light sensor, and screen usage (lock/unlock). Example proxies are listed in Figure 2 (depression) and Figure 3 (PD). For depression these include actions and behaviors indicating changes in sleep, physical and social activity, and cognitive function, while for PD useful biomarkers are found to relate to motor function and voice production.

These proxies form the basis for I+FLL, where patient-label supervision (from patients with known disease status) is used to induce a preliminary model, which is then refined through feature label supervision encoding relevant medical knowledge. As mentioned above, if labeled training examples are especially difficult to acquire, LSP enables the available data to be augmented with synthetic examples. The complete predictive modeling process is discussed in greater detail in the next two subsections of the paper.

Given passively-collected smartphone sensor data and associated TD diagnoses for a (small) set of patients, the goal is to learn a model capable of accurately predicting the probability that a new patient is suffering from the TD. The focus on depression and PD as brain disorders of interest leads to the following specific prediction targets: determine whether an individual: i.) is ‘depressed’ (PHQ-9 score ≥ 10) or ‘not depressed’ (PHQ-9 score < 10) [24], ii.) would be diagnosed with PD if examined by a specialist [e.g. 18,19].

To permit a quantitative problem formulation, we introduce data models for the two sources of learning-supervision:

- Labeled patient cohort model D_L_: given a training set of N patients, D_L_ is assembled by
  - representing each individual with feature vector **z** containing raw smartphone sensor data;
  - transforming **z** to disorder-relevant feature vector **x**∈X⊆ℜ^|F|^ (F = {f_1_,…, f_|F|_} is the condition-relevant feature set) through a map **z** → **x** which captures medical domain knowledge;
  - appending instance label y∈{−1,+1} to each patient (y=+1 for those with the target disorder), yielding D_L_ = {(**x**_1_, y_1_),…, (**x**_N_, y_N_)}.
- Labeled feature-set model D_F_: assume there is a subset of features F_L_⊆F that, according to domain knowledge, are associated with TD; suppose for convenience the features are binary (x_i_∈{0,1} and x_i_ = 1 if feature f_i_ is present); each feature f_i_∈F_L_ can be thought to possess a label u_i_∈{−1,+1}, with u_i_ = +1 if presence of f_i_ is linked to positive TD-state; importantly, true labels u_i_ are considered unknown – medical knowledge is used only to encode initial estimates u_0i_∈{−1,+1} for these labels; D_F_ is then defined as D_F_ = {(f_1_, u_1_),…, (f_l_, u_l_)}.

With these definitions in place, the TD-detection problem can be stated. Let Tar = {**z**_N+1_,…, **z**_N+T_} denote the target cohort of patients for whom TD-status is to be predicted (i.e. labels for these patients are not known). Use D_L_, D_F_, **u**_0_ = [u_01_,…, u_0l_]^T^, and map **z** → **x** to learn a model which accurately predicts TD-status y of any patient **z**∈Tar.

### B. Prediction Methodology

This subsection describes our baseline method for learning brain disorder-detection models from limited training data. If necessary, this core procedure can be augmented through the introduction of synthetic training examples; this option is presented in the next subsection. The baseline scheme consists of three main steps. First, raw feature vector **z** is transformed to representation **x, z** → **x**, by leveraging medical domain knowledge. Second, instance-labeled dataset D_L_ is used to induce prediction model **f**_L_: X → p_0_, where p_0_ is the ‘preliminary’ predicted probability that patient **x** has label y = +1 (i.e. has the target disorder). Let Tar_**x**_ = {**x**_N+1_,…, **x**_N+T_} denote the set of patients Tar when each is represented with condition-relevant feature vector **x** rather than raw features **z** (via feature mapping **z** → **x**). Applying **f**_L_ to unlabeled dataset Tar_**x**_ yields vector of predictions **p**_0_ = [p_0(N+1)_,…, p_0(N+T)_]^T^, one for each of the T patients in Tar_**x**_. However, if the number of labeled examples N used to learn **f**_L_ is small, predictions **p**_0_ may not be accurate. This observation motivates the third step in the TD detection strategy: refine predictions **p**_0_ → **p** = [p_N+1_,…, p_N+T_]^T^, with p_i_ the predicted probability **x**_i_∈ Tar_**x**_ has TD, by incorporating feature label dataset D_F_ and the initial label estimates **u**_0_.

Each step in the model-learning methodology is now described in greater detail.

#### Feature mapping z → x

Mapping raw features **z**, reflecting smartphone sensor outputs, to disorder-relevant features **x** typically provides important benefits, including improved prediction accuracy, increased interpretability, and enhanced compatibility with clinical objectives (e.g. therapy). Because feature vector **x** is disorder-specific, the **z** → **x** transformation is constructed separately for depression and PD.

For depression, the map draws on established DSM-5 symptoms [17]. A general summary of symptoms and proxies involved is listed in Table 1.

**Table 1.** Feature map: depression.

| DSM-5 Symptom | Smartphone Proxy Features |
| --- | --- |
| sleep change | estimated sleep-time pattern (light sensor, lock/unlock) |
| diminished concentration | phone usage patterns (lock/unlock) |
| diminished activity: physical | mobility/activity patterns (GPS, accelerometer) |
| diminished activity: social | estimated conversation patterns (microphone) |

The particular **z** → **x** map implemented in this paper transforms raw smartphone sensor readings into 15 psychological features (other instantiations are possible [23]):

- sleep change: depression-relevant features are the mean and variance of daily ‘go-to-bed time’, ‘wake-up time’, and their difference, estimated using the phone’s ambient light sensor together with detected screen lock/unlock events [4];
- diminished concentration: proxy features are mean and variance of daily numbers of screen locks/unlocks during time periods commonly devoted to cognitively-demanding tasks (e.g. classes, study), estimated from time-series data for phone locks and unlocks;
- diminished interest in physical activities: candidate relevant features are i.) mean and variance of daily travel distance, and ii.) entropy of the activity state (classified as ‘stationary’, ‘walking’, ‘running’, or ‘unknown’), estimated via GPS coordinates and accelerometer output, respectively [4];
- diminished interest in social activities: proxy features are mean and variance of the daily frequency and duration of conversation activity, estimated using microphone data [4].

The **z** → **x** map for PD captures current understanding of clinical presentation of the disease [18,19]; see Table 2 for an overview of symptom categories and proxies. The physiological features employed in this paper are summarized below (the full list of 22 features which make up the **z** → **x** map are reported in [25]):

**Table 2.** Feature map: PD.

| PD Symptom | Smartphone Proxy Features |
| --- | --- |
| voice production | speech characteristics (microphone) |
| impaired mobility | movement patterns (accelerometer, GPS) |

- speech: PD-relevant features are attributes of the patient’s voice, including statistical quantities (e.g. fundamental frequency mean and maximum, amplitude variance) and information-theoretic/complexity measures (e.g. signal fractal dimension, pitch entropy), all estimated from recordings made with a phone’s microphone (comprising 22 features in total);
- movement: proxy features quantify the statistics associated with the patient’s movement (e.g. tremor, balance, rigidity, fatigue), estimated via accelerometer and GPS data [25] (note that movement features are not used in the present investigation).

#### Preliminary prediction x → p_0_

The preliminary predictions **p**_0_ are formed by learning a model **f**_L_: X → p_0_ from instance-labeled dataset D_L_ and applying **f**_L_ to unlabeled patient dataset Tar_**x**_. This computation gives vector of predictions **p**_0_ = [p_0(N+1)_,…, p_0(N+T)_]^T^, one for each patient in Tar_**x**_. The predictive modeling is straightforward using any algorithm which permits statistically-efficient supervised learning. Here we adopt the random forest (RF) model for **f**_L_, with 1000 trees and default hyperparameter values [20] (varying the tree number between 200 and 2000 has little effect).

If the number of labeled training examples N from which model **f**_L_ is induced is too small, predictions **p**_0_ may not be sufficiently accurate. This possibility is addressed with the third step in the proposed learning method.

#### Prediction refinement p_0_ → p

One way to improve predictions **p**_0_ is to exploit two sources of information which are independent of D_L_: i.) characteristics of the underlying structure of the data, revealed through unsupervised clustering of unlabeled dataset Tar_**x**_ [26], ii.) medical knowledge, as encoded in feature label dataset D_F_. It is easy to see that integrating medical knowledge into the disease prediction process may yield benefits. The utility of unlabeled patient dataset Tar_**x**_ derives from the assumption that if patients **x**_k_ and **x**_l_ are ‘similar’ then they should possess similar labels. This, in turn, suggests that patient-similarity inferred from unlabeled data could be of value for improving the predictions **p**_0_.

We now present a computational methodology for using unlabeled patient data and medical knowledge to refine preliminary predictions **p**_0_. Recall that D_L_ = {(**x**_1_, y_1_),…, (**x**_N_, y_N_)} and Tar_**x**_ = {**x**_N+1_,…, **x**_N+T_} are labeled and unlabeled patient datasets, respectively. Consider the task of computing predictions **p** = [p_N+1_,…, p_N+T_]^T^ and **u**_f_ = [u_f1_,…, u_fL_]^T^, where p_i_, u_fi_ are the predicted probabilities that patient **x**_i_∈Tar_**x**_ has label y_i_ = +1 and feature f_i_∈F_L_ has label u_i_ = +1, respectively. These predictions are facilitated by modeling the relationships between the patients in Tar_**x**_ and their attributes with a bipartite graph G of instances and features, where an edge is placed between labeled feature f_j_ and patient **x**_i_ if and only if f_j_ is ‘present’ in **x**_i_ (i.e., (**x**_i_)_j_ = 1 [21]).

The desired predictions **p** are obtained by solving the optimization

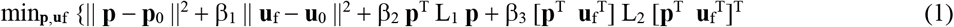

subject to the constraints p_i_∈[0,1] and u_fi_∈[0,1]. In (1), L_1_ = I−C with I the identity matrix and C a similarity matrix computed via ensemble clustering on Tar_**x**_ (i.e. element C_kl_ of C equals the number of times patients **x**_k_ and **x**_l_ are assigned to the same cluster by the ensemble), L_2_ is the Laplacian for bipartite patient-feature graph G, and β_1_, β_2_, β_3_ are nonnegative weights (see [21,26] for a description of the calculation and properties of Laplacian matrices L_1_ and L_2_).

It is easy to show that L_1_ measures similarity between pairs of patients **x**_k_ and **x**_l_ and L_2_ measures similarity between patients and their (labeled) features. Thus (1) can be seen to balance four objectives: i.) patient-label predictions **p** should be close to **p**_0_, ii.) feature label predictions **u**_f_ should tend to agree with **u**_0_, iii.) similar patients ought to have similar labels, iv.) patient labels should tend to match the labels of their features. The values of hyperparameters β_1_, β_2_, β_3_ establish the relative importance of these goals.

We summarize the preceding discussion by sketching an algorithm for predicting the probability that each patient in dataset Tar has the TD of interest. As the scheme learns from both instance labels and feature labels, the algorithm is called Algorithm I+FLL.

##### Algorithm I+FLL (Instance + Feature Label Learning)

Given D_L_, D_F_, **u**_0_, mapping **z** → **x**, and cohort Tar = {**z**_N+1_,…, **z**_N+T_} for whom TD-status predictions are to be made:

1. Transform patient cohort model Tar = {**z**_N+1_,…, **z**_N+T_} to Tar_**x**_ = {**x**_N+1_,…, **x**_N+T_} with the mapping **z** → **x**.
2. Form preliminary predictions **p**_0_ = [p_0(N+1)_,…, p_0(N+T)_]^T^ using model **f**_L_: X → p_0_ learned from instance-labeled dataset D_L_.
3. Compute (final) predictions **p** = [p_N+1_,…, p_N+T_]^T^ by solving (1) with convex programming (e.g. block coordinate descent [27]).

Remark: because (1) is convex this optimization can be completed efficiently and Algorithm I+FLL can be applied to large-scale problems.

### C. Learning from Synthetic Patients

In some applications, the labeled training data is extremely limited and/or class-imbalanced, and this is an obstacle to effective learning. This situation arises, for example, because it is often difficult to collect smartphone data traces for individuals with known status for the TD of interest, and because typical datasets have far fewer ‘cases’ (patients with the TD) than ‘controls’ (unaffected patients). We hypothesize that, if sufficiently-realistic ‘synthetic’ patient vectors could be acquired, especially for the minority-class, adding these vectors to the training set could improve the predictive modeling process. Specifically, it may be possible to increase the predictive accuracy of Algorithm I+FLL by generating synthetic patient vectors, combining these vectors with real labeled examples, and training Algorithm I+FLL on this larger ‘hybrid’ dataset. The basic aspects of this learning from synthetic patients (LSP) method are now described. (See [23,22] for a more thorough exposition.)

Previous studies have found that, to be useful for improving learning performance, synthetic examples should be *realistic* and *diverse* [28,29,23]. We integrate two complementary mechanisms to create such synthetic patients.

- Realism is achieved by using adversarial learning to produce synthetic patient feature vectors (SPs) that are indistinguishable from those of real patients (RPs) [22,23] and by working directly in the original patient feature space.
- Diversity is obtained by appropriately injecting randomness into SP construction, with an emphasis on outputting SPs which are well-separated in feature-space [28,22,23].

As is demonstrated empirically in subsequent sections, the resulting SPs are useful and convenient for downstream analysis (e.g. predictive modeling) and also intuitively-interpretable by clinicians.

The iterative algorithm used to build synthetic patients which are both realistic and diverse is illustrated in Figure 4. Informally, at each iteration, the realism of current SPs is increased by adapting their feature vectors so as to reduce the ability of a well-trained classifier to distinguished them from RPs. SP-diversity is encouraged primarily through the use of two stochastic processes: random initialization of SP ‘seed’ vectors and random modification of the adaptation procedure aimed at increasing SP-realism.

**Figure 4.**
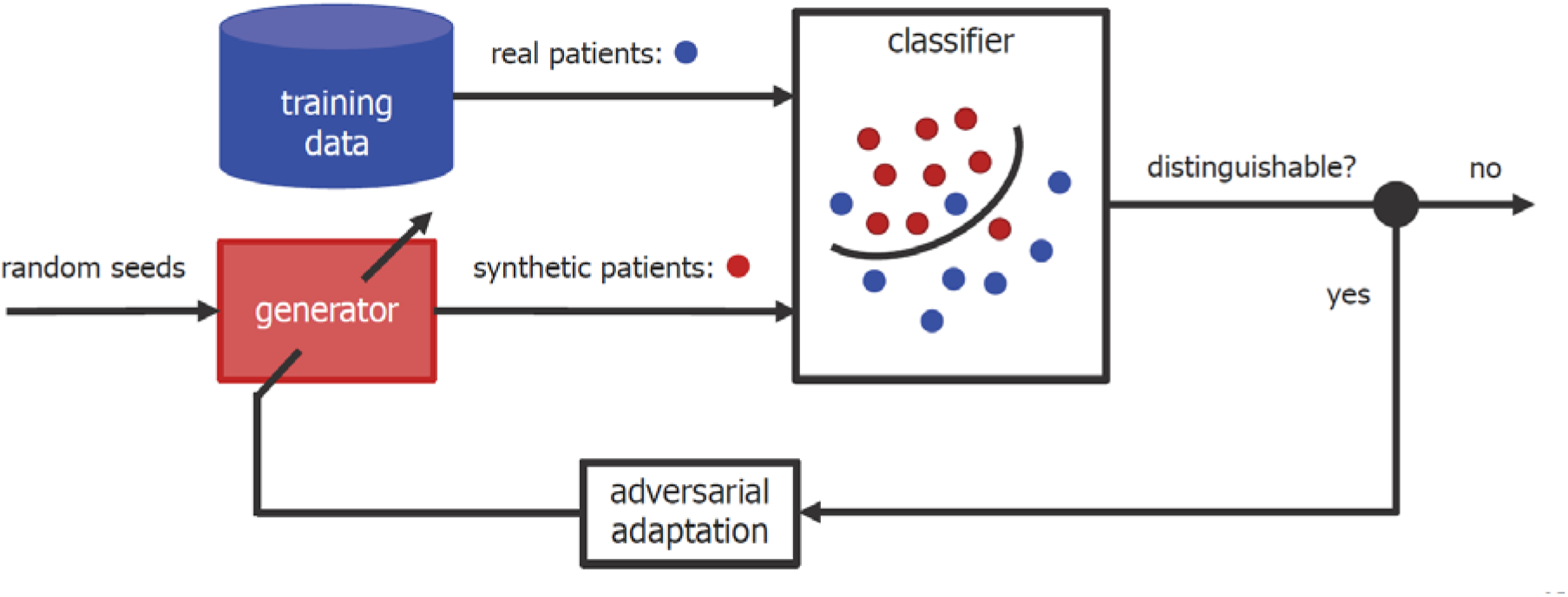
Schematic of adversarial synthetic patient generation.

This basic idea is formalized with the SP generation and learning algorithm specified below. The main focus is applications with very few positive-class training examples, as this is common in smartphone-based health monitoring. However, as delineated below, synthetic patients corresponding to both positive-class and negative-class RPs can be created if needed.

Algorithm LSP (Learning from Synthetic Patients)

- Input: a set of real patients RP = {**x**_1_,…, **x**_R_} drawn from the minority-class training set, the desired number of synthetic patients S, a threshold ε∈[0.5,1] for real v. synthetic patient distinguishability, and hyperparameters β_4_≥0, β_5_≥0, λ∈[0,1].
- Initialization: draw random ‘seed’ synthetic patients SP = {**s**_1_,…, **s**_S_} from a high-entropy distribution D_SP_ with feature-space support similar to RP.
- Synthetic patient generation process: iterate the following two steps until the sets RP and SP cannot be distinguished (classifier area under ROC curve < ε):
  1. Adaptation step: solve min_**w**_ max_{**a**1,,…**a**S}_ { Σ_i=1,R_ loss(+1, **w**^T^ **x**_i_) + Σ_j=1,S_ loss(−1, **w**^T^ (**s**_j_ + **a**_j_)) + β_4_ ‖ **w** ‖^2^ − β_5_ Σ_j=1,S_ ‖ **a**_j_ ‖ }
  2. Refinement step: for each **a**_j_  **0** (adaptation step encourages unnecessary **a**_j_ to be ‘zeroed-out’)

sample real patient **x** uniformly at random;
sample δ∈[0,1) uniformly at random;
refine **s**_j,new_ = **s**_j_ + λ **a**_j_ + (1−λ) δ (**x** − **s**_j_).
  3. Learning from synthetic patients:

Apply Algorithm I+FLL to the hybrid dataset formed by adding synthetic patients SP to the original (real) patient training dataset.

If the majority class training examples are very limited as well, synthetic patients may be created for both the positive class (SP_+_) and negative class (SP_−_), and then SP = SP_+_ ⋃ SP_−_ can be exploited for model training.

## 3. Experiment One: Detecting Depression in College Students

We evaluated the performance of Algorithm I+FLL and Algorithm LSP for the task of detecting and monitoring depression through experiments with the StudentLife [4] and DemonicSalmon [9] datasets. More specifically, this section: i.) defines the learning tasks of interest, ii.) offers a concise overview of the StudentLife and DemonicSalmon datasets, iii.) assesses detection accuracies of Algorithms I+FLL (Section 3.B) and LSP (Section 3.C) and compares their performance with strong benchmarks [5,8], iv.) reports which smartphone-derived proxy features have predictive power and provides clinical interpretations of these features, v.) investigates the efficacy of models learned with Algorithm I+FLL for monitor patients and identifying depression trends (Section 3.B).

### A. Setup

The depression detection and monitoring experiments are conducted using two publicly-available datasets: Student-Life [4] and DemonicSalmon [9]. The portion of StudentLife used here is a set of de-identified, passively-collected smartphone sensor outputs and survey results which:

- were gathered for 40 Dartmouth College students over a ten-week period;
- characterize each student’s location traces (GPS latitude-longitude coordinates), communication activity (number and duration of conversations, inferred from microphone data), physical activity level (‘stationary’, ‘walking’, ‘running’, or ‘unknown’, estimated via accelerometers), ambient light level (from phone light sensor), and phone usage (screen lock/unlock);
- contain self-reported PHQ-9 depression scores for each student, obtained at beginning and end of the ten-week study period (PHQ-9 is a validated depression appraisal tool [24]).

Similarly, DemonicSalmon includes de-identified, passively-collected smartphone sensor outputs and survey results which:

- were gathered for 68 University of Virginia students over a two-week period;
- characterize each student’s communications (the frequency of call and SMS conversations, inferred from phone data) and physical activity level (reflected in raw accelerometer data);
- contain daily self-reported PANAS positive/negative affect scores (PANAS is a validated affect assessment tool [30]).

The main goal of this section is to use Algorithm I+FLL to learn smartphone-based prediction models which enable accurate i.) inference of an individual’s depression state (‘depressed’ if PHQ-9 score ≥ 10, ‘not depressed’ if PHQ-9 score < 10 [24]), and ii.) monitoring of patient depression trend (‘worsening’ if ΔPHQ-9 ≥ 0, ‘improving’ if ΔPHQ-9 < 0) or positive/negative affect trend (change in PANAS score [30]). The primary measure of prediction quality is area under the ROC curve (AUC), estimated through leave-one-out cross-validation (together with statistical significance of performance differences [31]). (Pearson correlation, and its statistical significance [20], is also computed to illuminate the predictability of rapidly-changing affect trends.)

To permit an objective, quantitative assessment of the performance of Algorithm I+FLL, we compare the accuracy of its predictions to those of two state-of-the-art benchmark models as well as a simplified version of the algorithm. The four depression-detection strategies implemented in the experiments are:

- ‘I+FLL’: Algorithm I+FLL deployed with 15 psychological features (see above), where preliminary predictions are made with an RF classifier trained on D_L_, prediction refinement is based upon two feature labels with initial estimates u_0,significant_sleep_change_ = +1, u_0,significant_phone_lock/unlock_ = +1, and β_1_ = β_2_ = β_3_ = 0.3;
- ‘I+FLL w/o FL’: Algorithm I+FLL as above, except with no feature label information (i.e., β_1_ = β_3_ = 0);
- ‘canzian’: support vector machine (SVM) classifier with carefully-engineered, mobility-oriented features [5];
- ‘wang’: state-of-the-art depression prediction model built using a logistic regression classifier with LASSO feature selection [8].

We also briefly examined standard supervised learning for this task, specifically SVMs and L2-regularized logistic regression without feature-engineering [20], but these models performed poorly (illustrating the difficulties experienced with conventional machine learning when training data is limited).

### B. Results: Detecting and Monitoring Depression with I+FLL

We begin by applying Algorithm I+FLL to the task of predicting depression state (‘depressed’ if PHQ-9 ≥ 10 or ‘not depressed’ if PHQ-9 < 10) for the 40 college students that make up our StudentLife cohort [4]. Observe that predicting depression status in real-time using passively-collected data is central to many clinical and research objectives (e.g. delivering timely and effective intervention). The experiments compare the utility of models learned via Algorithm I+FLL with strong benchmark models (see above), depression-screening tools currently used in clinical practice, and diagnostic accuracy of treating physicians. In addition, predictive power of individual psychological feature categories (e.g. mobility-based) is appraised.

The results of the depression-detection experiments are shown in Figures 5 and 6. It is seen from Figure 5 that a prediction model learned using Algorithm I+FLL (magenta bar) significantly outperforms benchmark models (AUC = 0.94 with p < 0.01 for AUC differences). Figure 6 reveals that, while combining psychological features from distinct DSM-5 symptom categories works best, some categories (social activity, sleep) enable good predictions when used in isolation.

**Figure 5.**
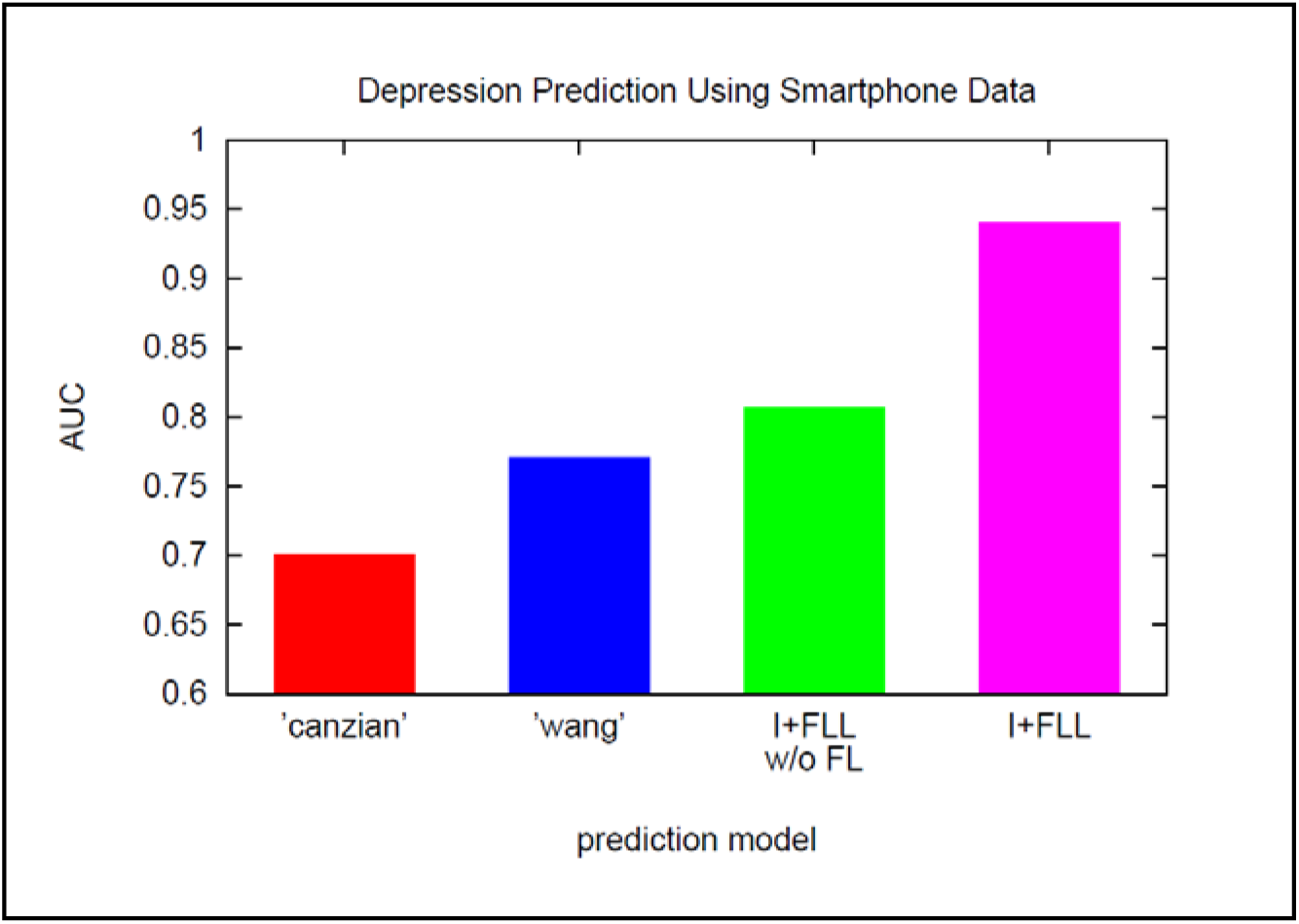
Depression-detection accuracy (AUC) of Algorithm I+FLL (magenta) and three benchmark models: ‘canzian’ [5] (red), ‘wang’ [8] (blue), and Algorithm I+FLL without feature label information (green).

**Figure 6.**
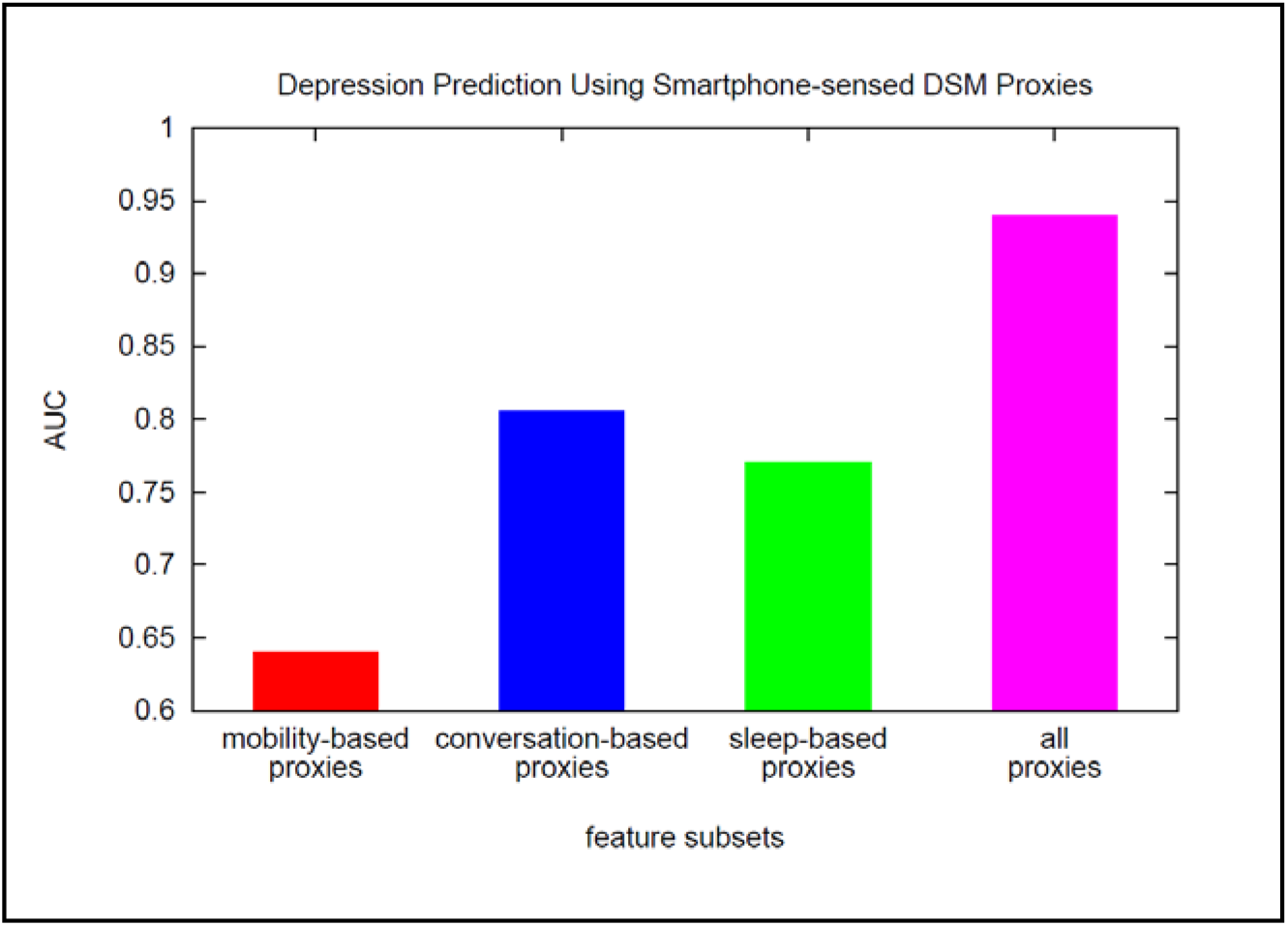
Depression-detection accuracy (AUC) of Algorithm I+FLL when using mobility proxies (red), conversation proxies (blue), sleep proxies (green), and all available features (magenta).

Also promising is a comparison of the depression-detection accuracy of Algorithm I+FLL, which requires only passive smartphone data, to that of high-quality depression-screening tools and diagnoses made by treating physicians. In this test, the evaluated depression-screening survey tools include PHQ-9, WBI-5, and HADS [32], diagnostic accuracy of physicians is as reported in [32], and ground-truth depression labels are assigned through Structured Clinical Interview for DSM-IV. The results of the study are as follows:

- screening tools: PHQ-9 AUC = 0.95, WBI AUC = 0.91, and HADS AUC = 0.89;
- treating physician: AUC estimated to be < 0.85;
- Algorithm I+FLL: AUC = 0.94.

Thus Algorithm I+FLL, which has access to only passively-collected smartphone data, outperforms HADS, WBI-5, and physicians, and is comparable to PHQ-9.

Knowing which features captured by smartphone sensors are predictive of depression is of value in various clinical applications. Feature predictive power was investigated using both forward- and backward-stepwise analyses [20]. Features possessing predictive power for depression include:

- conversation frequency and duration (measuring diminished interest/ pleasure in social activity);
- sleep-time variance (a proxy for sleep change);
- travel variance and physical-activity entropy (measuring diminished interest/pleasure in physical activity);
- phone lock/unlock (reflecting diminished ability to concentrate).

It is also of interest to explore the utility of Algorithm I+FLL for smartphone-based patient monitoring, for example to distinguish between improving and worsening depression trends. Toward that end, the next set of experiments are focused on using smartphone sensor traces to accomplish two tasks: i.) recognize improving (ΔPHQ-9 < 0) v. worsening (ΔPHQ-9 ≥ 0) depression trends over a ten-week period for 40 college students in the StudentLife dataset, ii.) detect daily changes in positive/negative affect (PANAS score) for 68 college students in the DemonicSalmon dataset. These task definitions are motivated by clinical and prediction-oriented considerations. For instance, detecting improving and worsening depression in real-time using passively-collected data has the potential to facilitate timely intervention and improve overall patient care [e.g. 33]. Moreover, successfully monitoring the depression of distinct cohorts over widely-varying time-horizons (monthly v. daily) would indicate that the proposed methodology enjoys desirable generalization properties.

In more detail, the monitoring tasks aim to:

- evaluate the capability of models learned via Algorithm I+FLL to distinguish between improving and worsening depression trends in the *low frequency* StudentLife dataset, where trends are captured by PHQ-9 assessments made for each student at the beginning and end of the ten-week study period;
- investigate the predictability of *high frequency* changes in positive/negative affect, as measured by daily self-reported PANAS scores for DemonicSalmon students, by computing correlations between these scores and the outputs of smartphone accelerometers (finding correlations between fine-grained affect changes and smartphone accelerometer readings is an important step toward building models capable of reliably predicting these changes [20]).

The results of the depression-monitoring experiments are shown in Figure 7 and Table 3. It can be seen from Figure 7 that trend-prediction models learned using Algorithm I+FLL (magenta bar) provide accurate monitoring, enabling improving/worsening depression to be distinguished with AUC = 0.89. Furthermore feature label learning is found to be a key ingredient in deriving good models (compare the magenta and green bars). Table 3 reveals that significant correlations exist in DemonicSalmon between daily trends in positive/negative affect (PANAS) and smartphone sensor data such as accelerometer outputs. Furthermore, the signs of the correlations agree with psychological consensus and their magnitudes appear sufficient to support predictive modeling; future work will examine this prospect in greater depth.

**Table 3.** Correlation Between Affect and Accelerometer Output.

| Affect Valence | Number of Students | Correlation with Mean of Accelerometer Norm | P-value | Correlation with Variance of Accelerometer Norm | P-value |
| --- | --- | --- | --- | --- | --- |
| positive affect | 68 | 0.1693 | 0.0021 | 0.2334 | 0.00002 |
| negative affect | 68 | -0.1488 | 0.0070 | -0.1753 | 0.0015 |

**Figure 7.**
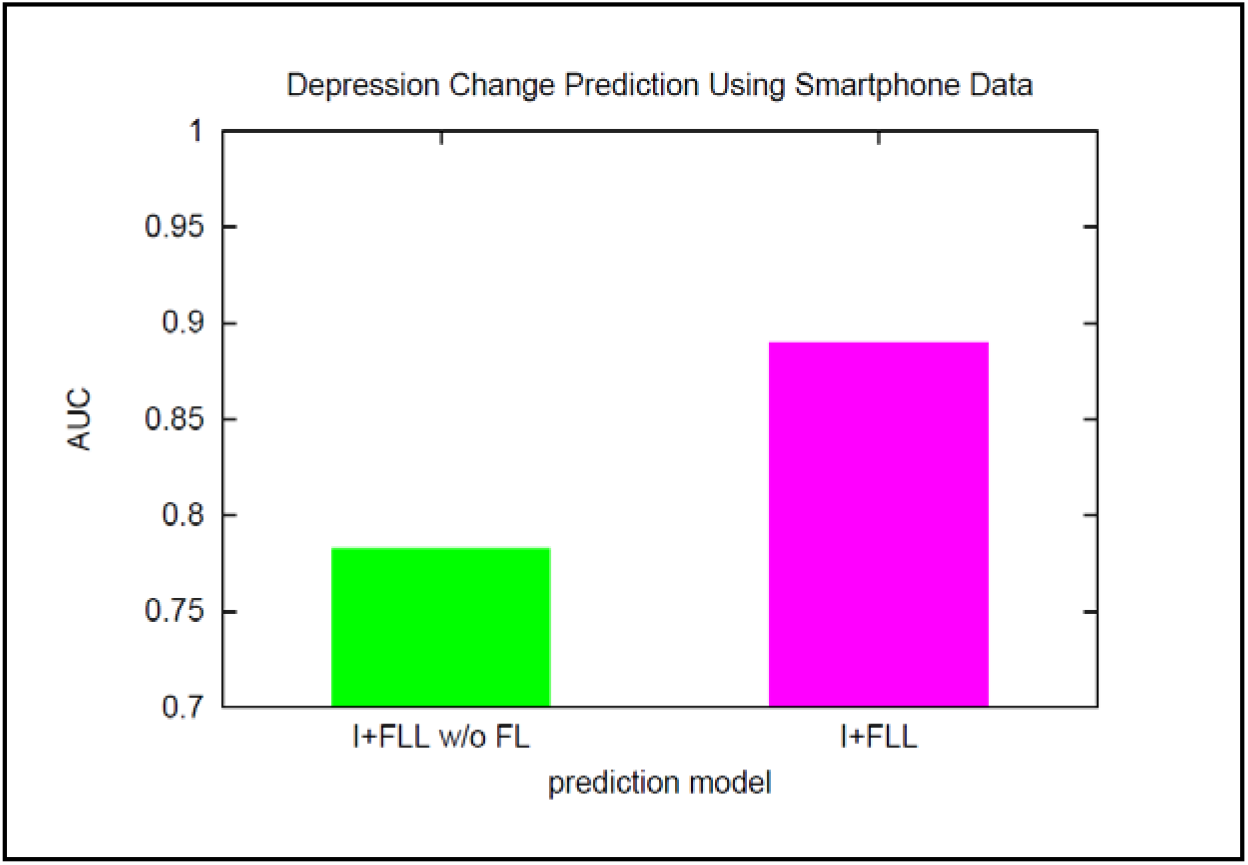
Depression monitoring accuracy (AUC) of Algorithm I+FLL (magenta) and Algorithm I+FLL without feature label information (green).

### C. Results: Enhancing Depression Detection via LSP

Observe that the StudentLife dataset contains a limited number of labeled examples from which to learn depression-detection models. The cohort studied here is composed of 40 students, 17 of whom are depressed, so models learned within a leave-one-out cross-validation protocol have access to only 39 examples. As discussed in Section 2.C, very small training sets represents a critical obstacle to effective learning, motivating study of the possibility to leverage ‘synthetic’ examples to boost predictive performance.

Consider the task of using Algorithm LSP to predict depression status (‘depressed’ if PHQ-9 ≥ 10, ‘not depressed’ if PHQ-9 < 10) for the 40 individuals in StudentLife [4]. To quantify the utility of Algorithm LSP in this reduced-data setting, we compare its predictions to those of the best-performing tools and methods tested thus far:

- ‘wang’: state-of-the-art depression prediction model built with logistic regression and LASSO feature selection [8];
- ‘PHQ-9’: the performance of the best available depression-screening survey tool (as reported in [32]);
- ‘I+FLL’: Algorithm I+FLL with hyperparameters defined as in Section 3.A;
- ‘I+FLL+LSP’: Algorithm LSP with I+FLL-hyperparameters defined as above, number of synthetic patients S = 20, distinguishability threshold ε = 0.55, β_4_ = β_5_ = 1, λ = 0.5, loss(.) = squared-loss, and D_SP_ = uniform.

The experimental results are displayed in Figure 8. Algorithm LSP (magenta bar) outperforms all the other methods, including PHQ-9 surveys, for depression-detection in the StudentLife dataset (AUC = 0.975). Therefore this analysis suggests that generating and learning from synthetic patients is an effective way to mitigate the challenges with very small training datasets.

**Figure 8.**
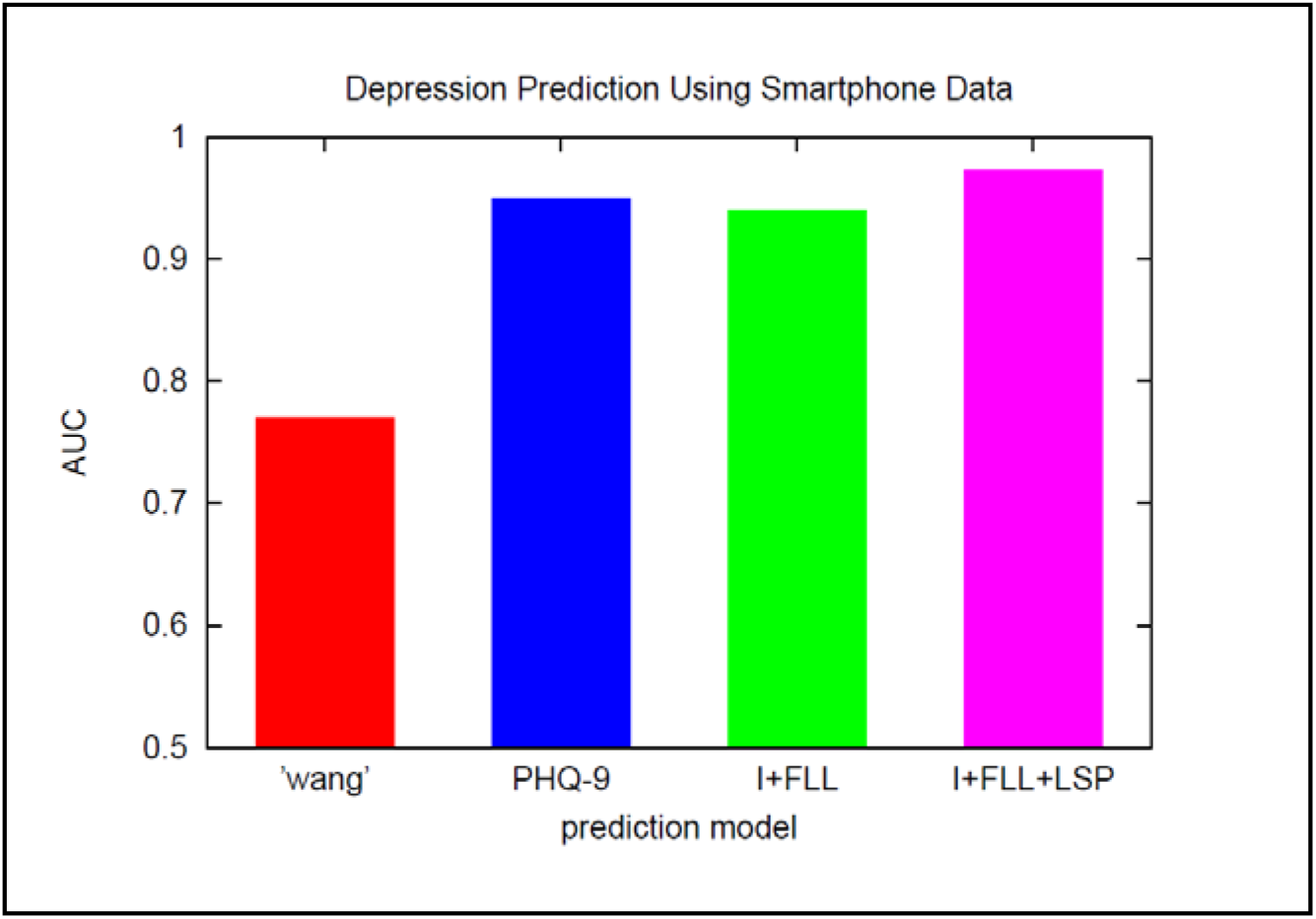
Depression-detection accuracy (AUC) of Algorithm LSP (magenta) and three alternative tools/methods: ‘wang’ [8] (red), ‘PHQ-9’ (blue), and Algorithm I+FLL (green).

## 4. Experiment Two: Detecting Parkinson’s Disease in Disparate Cohorts

We now evaluate the performance of Algorithm I+FLL for the task of detecting PD from smartphone recordings of a person’s voice. To enable robustness and generalizability of the approach to be investigated, two different datasets, corresponding to disparate patient cohorts, are used in the experiments: i.) a ‘US cohort’ of 31 individuals, 23 of whom have PD [10], ii.) a ‘Turkish cohort’ of 40 individuals, 20 of whom have PD [12]. The accuracy of Algorithm I+FLL is assessed by comparing its predictions with those of both strong benchmark models and established PD screening questionnaires. Additionally, we study the predictive power of distinct feature subsets obtained through different audio signal processing schemes.

### A. Experiments with US Cohort

The prediction task in the first set of experiments is to learn models which accurately determine whether a given individual has PD based upon brief smartphone recordings of the person’s voice. The ‘US cohort’ dataset [10] used in these experiments:

- is composed of 195 (de-identified) voice recordings for 31 individuals, 23 of whom have PD, with most recordings lasting for a few seconds; each voice recording is described by 22 features expected to be PD-relevant [10,25], including
  - 16 standard ‘statistical’ attributes (e.g. mean/maximum/minimum fundamental frequency, variation of fundamental frequency and signal amplitude, signal-to-noise ratio) – see [25] for details;
  - 6 information-theoretic/complexity measures (e.g. fractal dimension, pitch entropy, dynamical complexity) – see [25] for details;
  - contains recordings of PD patients in the early stages of the disease – for example, 26% have little or no functional disability (stage ≤ 1.5 on the Hoehn/Yahr scale [10]) and 35% have been diagnosed for ≤ 2yr;
- includes a label for each patient, ‘PD’ or ‘healthy’, assigned by PD specialists based upon thorough clinical examination (see [10] for details).

Predictive performance is measured with area under the ROC curve (AUC), estimated through leave-one-out cross-validation (together with statistical significance of performance differences [31]). Quantitative assessment of the accuracy of Algorithm I+FLL is facilitated through comparison with two state-of-the-art benchmark models as well as two simplified version of the algorithm. The five PD-detection strategies implemented in the experiment are:

- ‘I+FLL’: Algorithm I+FLL deployed with all 22 physiological features (see above and [25]), where preliminary predictions are made with an RF classifier trained on D_L_, prediction refinement is based upon two feature labels with initial estimates u_0,increased_pitch_entropy_ = +1, u_0,increased_signal/noise_ = −1, and β_1_ = β_2_ = β_3_ = 0.3;
- ‘I+FLL-statistical’: Algorithm I+FLL as above but using only 16 voice statistics as features (e.g. mean fundamental frequency, signal/noise ratio) [25];
- ‘I+FLL-information-theoretic’: Algorithm I+FLL as above but using only 6 information-theoretic and complexity quantities as features (e.g. pitch-entropy, fractal dimension of voice signal embedding) [25];
- ‘little’: state-of-the-art PD detection model built with an SVM classifier and extensive feature engineering [10];
- ‘psorakis’: relevance vector machine with sophisticated basis function and training sample selection [34].

Tests of standard learning methodologies (traditional SVMs, logistic regression) [20] for this task suggest these perform poorly, likely because the training dataset is small.

Results of the ‘US cohort’ PD-detection experiment are shown in Figure 9. It is seen that a prediction model learned using Algorithm I+FLL (cyan bar) significantly outperforms all benchmark models (AUC = 0.985 with p < 0.01 for AUC differences). Notice that the sets of statistical and information-theoretic/complexity features are predictive on their own and complementary when combined.

**Figure 9.**
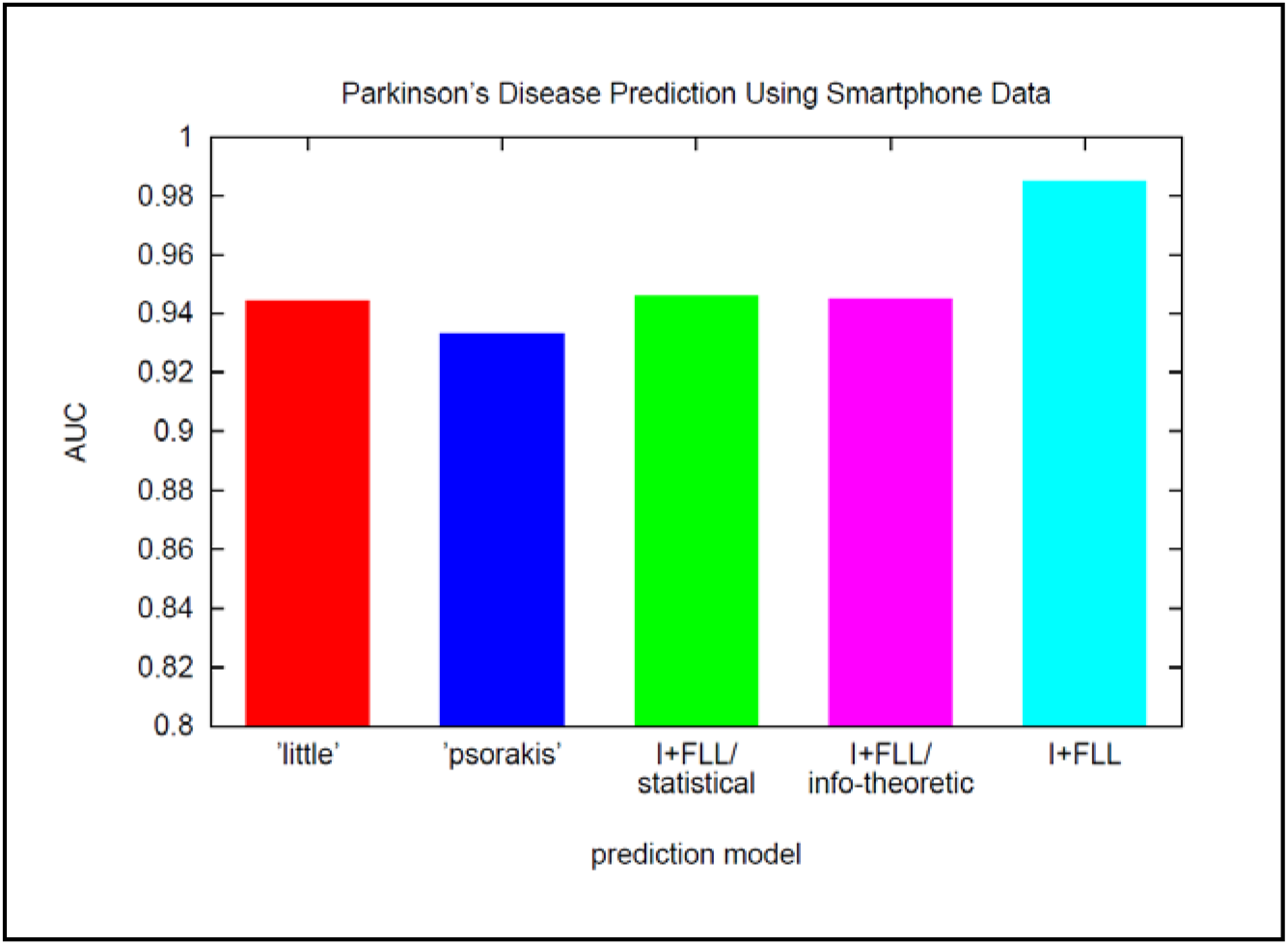
PD-detection accuracy (AUC) of Algorithm I+FLL with all features (cyan) and four benchmark models: ‘little’ [10] (red), ‘psorakis’ [34] (blue), Algorithm I+FLL with statistical features (green), and Algorithm I+FLL with information-theoretic/complexity features (magenta).

We also compared the PD-detection accuracy of Algorithm I+FLL, which requires only passive smartphone data, to that of high-quality PD-screening questionnaires. PD-screening tools evaluated in this test include NMS-Quest [35] and Tele-Quest [36], and ground-truth PD labels reflect diagnoses made by PD specialists based upon thorough clinical examination. Because [35,36] report sensitivity and specificity [20], rather than AUC, those are also used here. The results of the study are:

- Algorithm I+FLL: sensitivity = 0.945, specificity = 0.960;
- NMS-Quest [35]: sensitivity = 0.718, specificity = 0.885;
- Tele-Quest [36]: sensitivity = 0.890, specificity = 0.880.

Thus Algorithm I+FLL, processing only smartphone data, achieves better screening accuracy than labor-intensive, clinically-validated PD questionnaires.

### B. Experiments with Turkish Cohort

The next set of experiments also involves learning models which detect PD in individuals based upon smartphone recordings of the person’s voice. In this case, though, we apply Algorithm I+FLL with a cohort that is quite different than the ‘US cohort’ – the individuals speak another language and are culturally and ethnically distinct. These differences allow examination of the algorithm’s robustness and generalizability.

In greater detail, the ‘Turkish cohort’ [12] used in these experiments

- consists of 1040 (de-identified) voice recordings for 40 individuals, 20 of whom have PD, with most recordings lasting for a few seconds; each voice recording is described by 26 features which are expected to be PD-relevant [10,12,25] and are similar to the ‘statistical’ attributes summarized above for the ‘US cohort’ (see [25] for more details);
- contains recordings of PD patients in the early stages of the disease; for example, no patient has more than mild impairment and 40% have little or no functional disability (UPDRS assessment [12]); additionally, no patient has been diagnosed for > 6yr;
- includes a label for each patient, ‘PD’ or ‘healthy’, assigned by PD specialists based upon thorough clinical examination (see [12] for details);
- has more feature measurement variability than the ‘US cohort’ dataset, motivating a modest extension to Algorithm I+FLL to provide enhanced robustness to feature noise (see below).

Three versions of Algorithm I+FLL are implemented, corresponding to distinct ways of mitigating the presence of feature noise in the ‘Turkish cohort’:

- ‘naïve’: Algorithm I+FLL with raw smartphone features, i.e., no feature label learning or physiological features (essentially the classifier developed in [37]);
- ‘feature-averaging’: Algorithm I+FLL with feature-averaging – each individual is modeled using the mean of that person’s recording-level features;
- ‘probability-averaging’: Algorithm I+FLL is applied to voice recordings, and individual-level predictions are formed by averaging that person’s recording-level predicted probabilities.

The results of PD-detection experiments with the ‘Turkish cohort’ are displayed in Figure 10. As is clear, the prediction-averaging version of Algorithm I+FLL (green bar) is substantially more accurate than the other schemes (AUC = 0.973 with p < 0.01 for AUC differences). This outcome indicates (recording-level) prediction-averaging may be a valuable extension of Algorithm I+FLL when smartphone sensor readings are especially noisy. These results and those of the preceding subsection offer evidence of the robustness and generalizability of the proposed methodology across quite-different patient cohorts.

**Figure 10.**
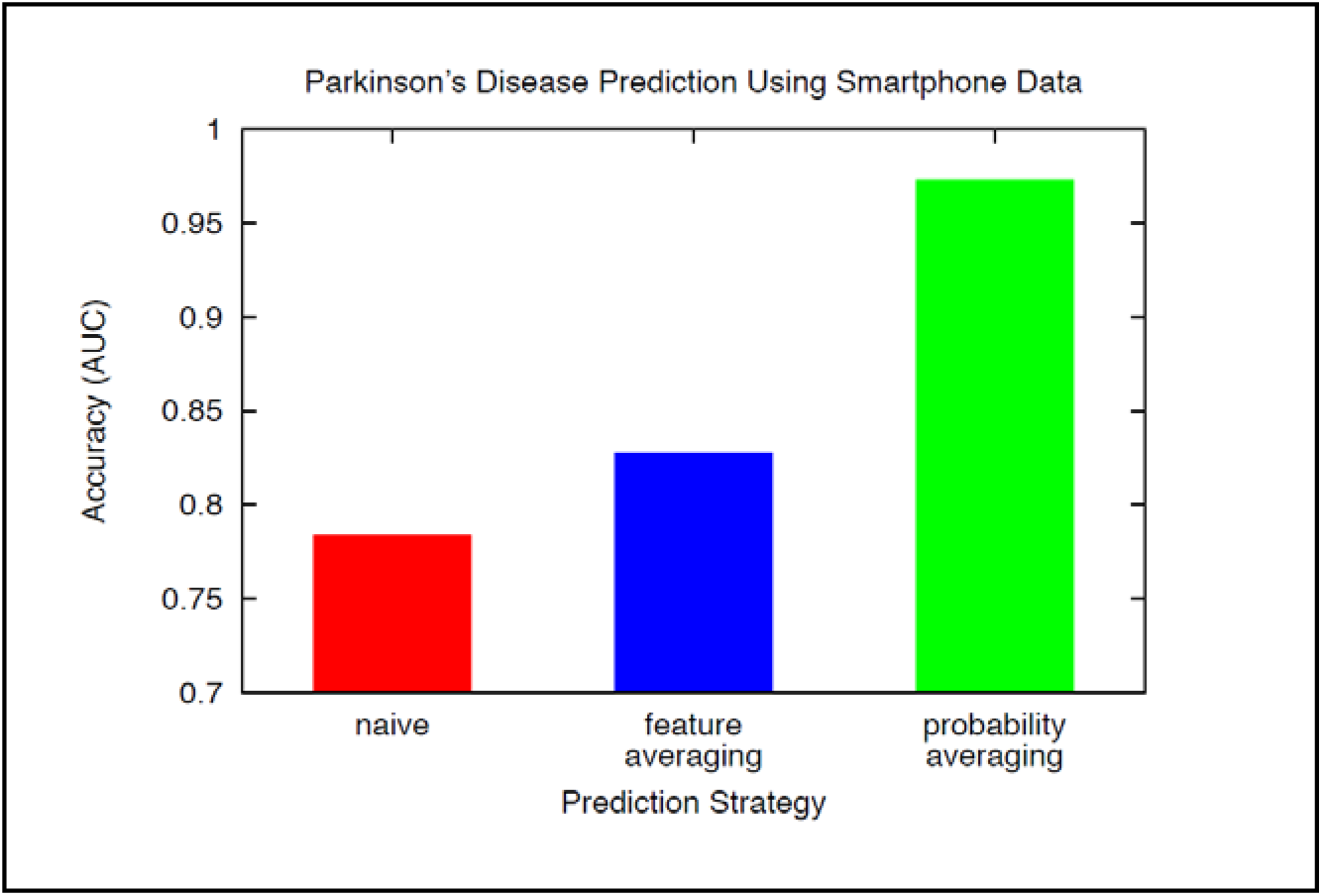
PD-detection accuracy (AUC) of three versions of Algorithm I+FLL: ‘naïve’ (red), prediction based upon patient’s average feature-vector (blue), and prediction by averaging patient’s recording-level model predictions (green).

## 5. Concluding Remarks

This paper presents a new method for leveraging passively-collected smartphone data and machine learning to detect and monitor brain disorders such as depression and Parkinson’s disease. Crucially, the algorithm is able learn accurate, clinically-compatible models from small numbers of labeled examples (i.e., smartphone users for whom sensor data has been gathered *and* disease status has been determined). Predictive modeling is achieved by learning from augmented training sets composed of both real and ‘synthetic’ patients. The proposed approach is shown to outperform state-of-the-art techniques in experiments involving disparate brain disorders and multiple patient datasets. Future work will include exploring the efficacy of the approach for other disorders.

## Data Availability

See citations in the manuscript for data availability.

